# Generalization Challenges in ECG Deep Learning: Insights from Dataset Characteristics and Attention Mechanism

**DOI:** 10.1101/2023.07.05.23292238

**Authors:** Zhaojing Huang, Sarisha MacLachlan, Leping Yu, Luis Fernando Herbozo Contreras, Nhan Duy Truong, Antônio Horta Ribeiro, Omid Kavehei

## Abstract

This research investigates the influence of dataset characteristics on the performance and generalization capabilities of deep learning models, on ECG data. The study evaluates multiple subsets of the TNMG dataset with varying levels of curated characteristics to assess their impact on model performance. Additionally, an attention mechanism is introduced to enhance model accuracy and generalization. The experimental results reveal that models trained on balanced subsets and incorporating the attention mechanism consistently outperform those trained on unbalanced data or without attention, emphasizing the critical importance of dataset balance and attention mechanism for achieving improved model performance.

Surprisingly, the largest ECG dataset, TNMG, proved less effective in generalization than smaller, curated subsets. The study demonstrates that a well-balanced and thoughtfully curated dataset, combined with the attention mechanism, can lead to competitive model performance, even with a significantly smaller size.

This research on ECG data underscores the critical importance of dataset curation, balance, and attention mechanisms in biomedical machine learning. It highlights that well-balanced, thoughtfully curated datasets with attention mechanisms can outperform larger, unbalanced datasets, challenging conventional notions and offering potential advancements in medical data analysis and patient care.

## 1 Introduction

Cardiac abnormalities are typically characterized as any deviation or alteration from an individual’s typical heart rate pattern that cannot be justified physiologically. They often indicate an underlying cardiac ailment and early identification can help prevent serious clinical conditions like heart failure or stroke [15].

Cardiac abnormalities are a common medical phenomenon affecting a significant portion of the global population. It is estimated that between 1.5 to 5% of people worldwide experience some form of abnormality [9]. There are, therefore, millions of people living with abnormalities which can impact their quality of life and overall health. The incidence of abnormalities tends to increase with age and is also influenced by comorbidities such as hypertension, diabetes, and pre-existing heart disease [5, 8]. This prevalence of abnormalities can strain healthcare systems worldwide due to the resources needed to diagnose and treat these conditions [15]. Moreover, untreated or undiagnosed abnormalities can lead to severe complications, further signifying the importance of early detection and proper management [12].

Multiple clinical methods are currently used to diagnose cardiac abnormalities, including examining the patient’s medical history, physical assessment, and specialized monitoring equipment. The electrocardiogram (ECG) is a widely utilized tool for measuring the heart’s electrical activity and was introduced by Waller in the early 1900s. It is considered essential in evaluating and diagnosing cardiovascular diseases [2]. The ECG offers a straightforward, low-cost, and non-intrusive approach to tracking heart signals. The most prevalent version is the standard 12-lead ECG (S12L-SCG); however, other types of ECGs use different numbers of leads for various purposes.

Physicians frequently encounter challenges in interpreting ECG recordings because of the intricacy, unclear causes of abnormalities, and their clinical associations. Depending solely on visual examination of ECG recordings can result in incorrect diagnosis and categorization, which can have life-threatening outcomes [6]. A high level of expertise is necessary to manually interpret ECGs [4]. Furthermore, there is variation in interpretation among different observers and even for the same observer over different readings, and the task is both monotonous and requires a lot of time even for highly experienced cardiologists [14]. Studies indicate that cardiologists can accurately recognize ECG abnormalities in the range of 53% to 96%, with as many as 33% of ECG readings containing some level of mistake, and up to 11% of cases leading to incorrect treatment [6]. As a result, the progress of digitalization in the healthcare sector has led to the introduction of computer-assisted ECG screening, which can aid medical professionals in identifying and diagnosing cardiovascular illnesses [6].

### 1.1 Related Work

Computer-aided diagnosis of cardiac conditions (CACD) can be a valuable resource for healthcare providers to obtain a second opinion and reduce diagnostic errors related to cardiac diagnosis [10]. Typically, CACD systems involve four primary stages: preprocessing of the ECG signal, heartbeat detection, extraction and selection of features, and construction of a classifier [1]. Despite advancements in computerized ECG interpretation, current systems still have a notable misdiagnosis rate, with an accuracy of 69.7% compared to 76.3% for cardiologists [6]. Traditional machine learning methods have made headway in tackling this issue, but they necessitate substantial expertise and pre-processing of the ECG signal for manual feature extraction [17]. Additionally, the considerable variation in wave morphology among patients and the existence of noise make achieving a high degree of accuracy challenging [17]. The development of deep neural networks (DNNs) through progress in machine learning and artificial intelligence has resulted in notable achievements in image and speech recognition and also holds promise for various healthcare and clinical applications.

Convolutional Neural Networks (CNNs), a type of deep learning algorithm, have effectively tackled complex image analysis tasks in various fields, including medical and non-medical applications [17]. These deep learning techniques entail supplying raw or minimally processed data into a network made up of numerous assembled mathematical equations or hidden layers, which enables the network to automatically extract, choose, and categorize features without the need for manual feature extraction [17]. Although DNNs have demonstrated good performance in classifying abnormalities, they are frequently confined to training on small, publically available datasets, need lengthy computation periods to tune the classifiers, and have not yet been verified in a clinical environment. By improving ECG interpretation, lowering misinterpretation, and minimising clinical treatment errors, a reliable, accurate, generalizable DNN with shown clinical performance has the potential to transform current practice by supporting clinicians in cardiac abnormality diagnosis.

Recent research has aimed to develop precise deep neural networks capable of accurately classifying cardiac abnormalities from ECG tracings. However, the generalization of these models has been given insufficient attention in some publications. Ribeiro *et al*. [19] constructed a DNN model that was trained on the most extensive 12 lead ECG dataset to date and achieved an impressive F1 score of over 80% and a specificity of more than 99% [19]. Nevertheless, this model has not been externally validated. The Hybrid Deep CNN Model proposed by Ullah [21] and the Deep-Learning-Based Framework introduced by Jamil [11] have both attained an impressive F1 score of 0.99 and above. However, neither of these models have been cross-validated on other datasets, and their ability to generalize to new data has not been evaluated.

The DNN model developed by Ribeiro *et al*. [19] boasts state-of-the-art performance after being trained on the largest ECG dataset. While it is commonly believed that larger datasets contribute to better model performance and improved generalization, this study aims to delve deeper into the dataset’s characteristics beyond just its size. By doing so, the research seeks to understand how these specific characteristics impact the model’s performance and generalization abilities. Zvuloni’s study revealed an intriguing pattern in the learning curves, as depicted in their recent work [27]. Initially, when working with a small dataset, traditional feature engineering (FE) techniques outperformed deep learning (DL) methods. However, as the dataset size increased, there was no significant discernible advantage between the performance of FE and DL approaches.

Moreover, this study explores the potential of attention mechanisms in enhancing model generalization to new cohorts. Attention mechanisms have garnered significant attention for their ability to improve model performance [3]. Through a series of experiments, assess how incorporating attention mechanisms influences the model’s capacity to generalize effectively. By doing so, we shed light on the dataset’s specific characteristics and the impact of attention mechanisms.

## 2 Background

### 2.1 DNN architecture

This study builds upon the model proposed by Ribeiro *et al*. [19]. Specifically, we utilize their unidimensional residual neural network architecture which includes a 1-D convolutional layer, followed by batch normalization and a ReLU activation function. The network then passes through four residual blocks before exiting with a dense layer, as depicted in Fig. 1a).

**Figure 1:**
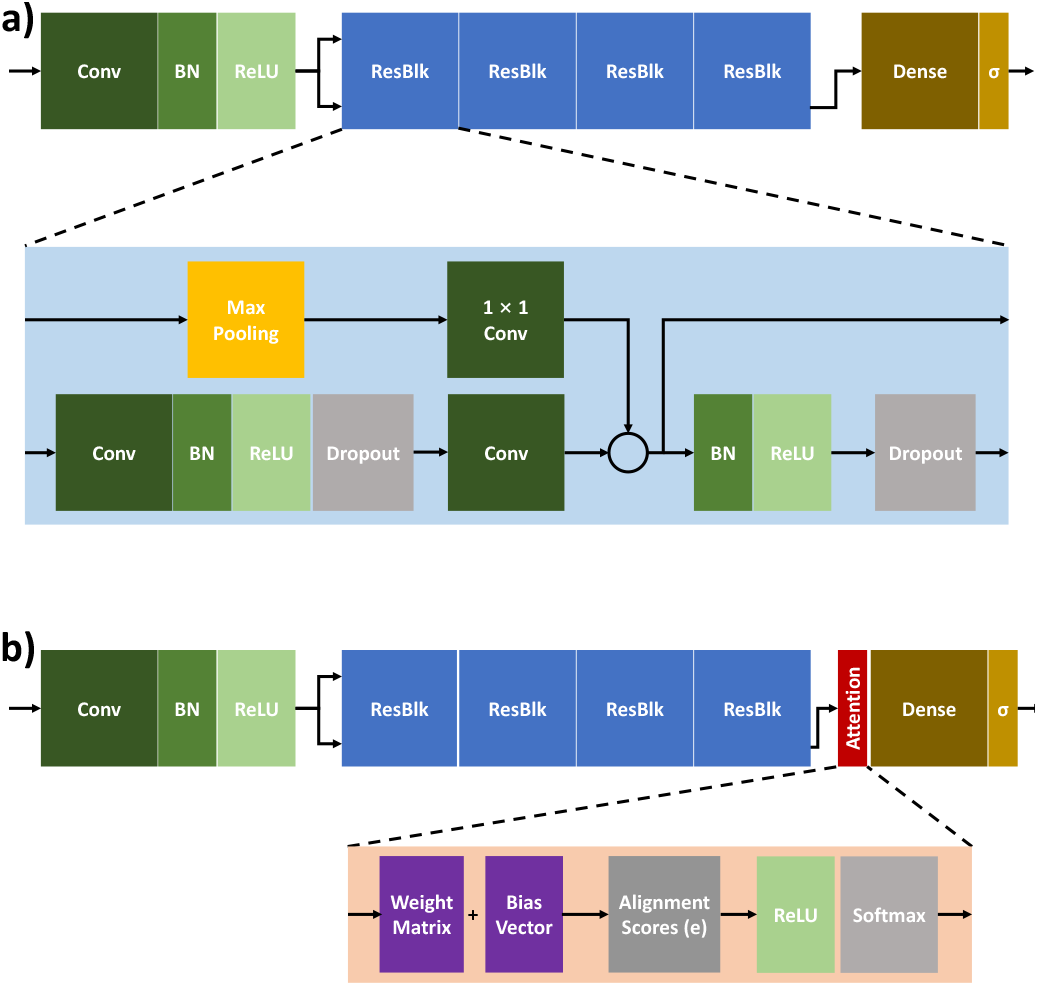
a) Ribeiro *et al*. [19] developed a comprehensive DNN model for abnormality classification, which utilized multiple ResBlks in a specific configuration. b) The attention layer added to the base model integrates both ReLU and Softmax mechanisms. It is positioned at the rear of the base architecture to study the effect of attention mechanisms on model performance and generalization.

### 2.2 Attention Layer

Bahdanau *et al*. [3] proposed an attention mechanism to address the bottleneck problem of fixed-length encoding vectors in sequence-to-sequence models. This problem is especially critical for longer or more complex sequences where the decoder’s access to input information is limited. The attention mechanism enables the decoder to selectively focus on relevant input parts, improving its access to information at each time step. The attention layer proposed in our model can be broken down into three main computational steps:

**Alignment scores** are computed by the attention mechanism’s alignment model, which takes encoded hidden states and the previous decoder output. This model is represented by a Eq. (1), typically a feedforward neural network.

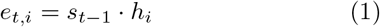

In the equation, *e*_*t,i*_ represents the alignment score between the decoder hidden state at time step *t−*1 (denoted by *s*_*t−*1_) and the encoder hidden state at time step *i* (denoted by *h*_*i*_). The alignment score is computed as the dot product between these hidden states and measures their relevance or similarity. This alignment score is used in the attention mechanism to determine the decoder’s focus on specific encoder hidden states during sequence generation tasks. The dot product captures the degree of alignment between the two hidden states and is commonly used in attention mechanisms to quantify their compatibility.

To calculate the **attention weights** (denoted by *a*_*t,i*_) as Eq. (2), the attention mechanism applies a softmax function to the alignment scores that were computed earlier.

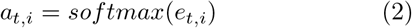

The attention mechanism generates a distinct **context vector** (denoted by *c*_*t*_) for the decoder at each time step as Eq. (3), which is determined by a weighted sum of all encoder hidden states.

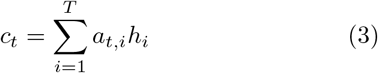

This simplified attention mechanism differs from the advanced self-attention used in paper “Attention is All You Need” [22]. Notable distinctions include a single attention head, absence of positional encoding, and a more straightforward architecture. This simplified approach may be better suited for specific tasks within a deep neural network model.

## 3 Methods

Using the state-of-the-art model presented by Ribeiro *et al*. [19] as the base model, this study aims to evaluate the cross-dataset generalization ability of the model. Additionally, the study investigates how training data characteristics impact the model’s generalization performance. Furthermore, the study proposes an attention mechanism as an effective solution to enhance the generalization ability of machine learning models.

### 3.1 Datasets

To evaluate the proposed method, this study primarily used the extensive ECG dataset collected by Tele-health Network Minas Gerais (TNMG) used by Ribeiro *et al*. [19]. Additionally, the China Physiological Signal Challenge 2018 (CPSC) dataset [13] and the ECG database from Shaoxing and Ningbo Hospitals (SNH) were employed as validation datasets.

#### Telehealth Network Minas Gerais (TNMG)

The Telehealth Network Minas Gerais (TNMG) dataset, cited in the reference by Ribeiro *et al*. [19], consists of 2,322,513 labeled S12L-ECG recordings with short duration, obtained from 1,676,384 unique patients. Between 2010 and 2016, the Telehealth Network of Minas Gerais, Brazil, gathered data using either a Tecnologia Eletronica Brasileira TEB ECGPC model or a Micromed Biotecnologia ErgoPC 13 model tele-electrocardiograph. Cardiologist annotations and the University of Glasgow ECG analysis program were used to label the recordings, resulting in a dataset containing a normal heart rhythm and six typical clinical abnormality categories. The recordings were sampled at 400 Hz, and zero-padding was used where necessary to ensure each of the 12 leads produced records of equal length, containing 4096 samples, which corresponds to approximately 10 seconds [19]. The six common clinical abnormality classes in the dataset are summarised in Table 1.

**Table 1:**
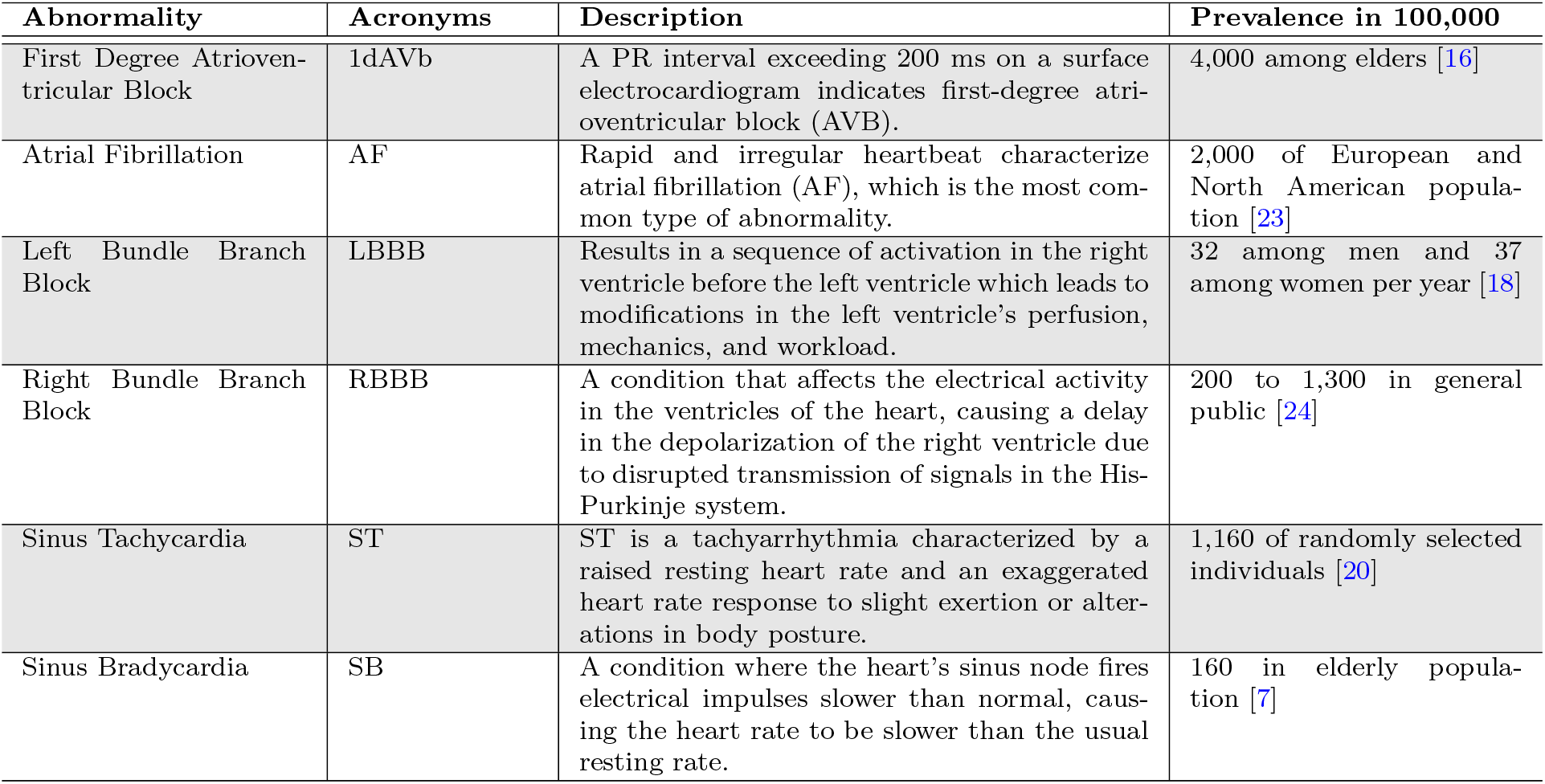
Classifications of Abnormality in TNMG Dataset and Incidents in General Population.

| Abnormality | Acronyms | Description | Prevalence in 100,000 |
| --- | --- | --- | --- |
| First Degree Atrioventricular Block | 1dAVb | A PR interval exceeding 200 ms on a surface electrocardiogram indicates first-degree atrioventricular block (AVB). | 4,000 among elders [16] |
| Atrial Fibrillation | AF | Rapid and irregular heartbeat characterize atrial fibrillation (AF), which is the most common type of abnormality. | 2,000 of European and North American population [23] |
| Left Bundle Branch Block | LBBB | Results in a sequence of activation in the right ventricle before the left ventricle which leads to modifications in the left ventricle’s perfusion, mechanics, and workload. | 32 among men and 37 among women per year [18] |
| Right Bundle Branch Block | RBBB | A condition that affects the electrical activity in the ventricles of the heart, causing a delay in the depolarization of the right ventricle due to disrupted transmission of signals in the His-Purkinje system. | 200 to 1,300 in general public [24] |
| Sinus Tachycardia | ST | ST is a tachyarrhythmia characterized by a raised resting heart rate and an exaggerated heart rate response to slight exertion or alterations in body posture. | 1,160 of randomly selected individuals [20] |
| Sinus Bradycardia | SB | A condition where the heart’s sinus node fires electrical impulses slower than normal, causing the heart rate to be slower than the usual resting rate. | 160 in elderly population [7] |

This study utilized 1,048,575 distinct ECG 12-lead tracings from the TNMG dataset as part of its experimentation. The distribution of the dataset is depicted in Figures 2a).

**Figure 2:**
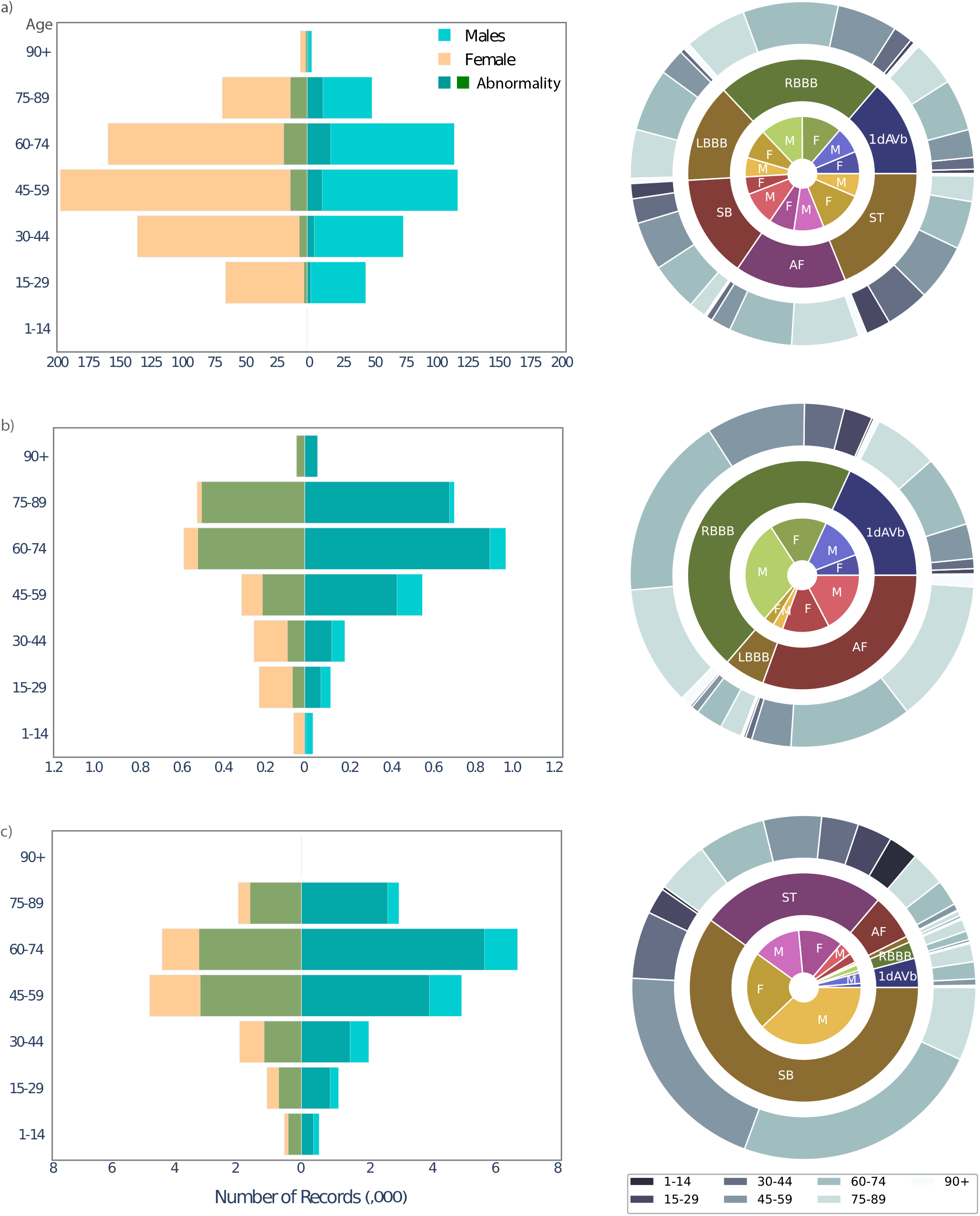
a) The TNMG dataset presents organized data for different age groups and genders. In this visualization, a darker bar represents the prevalence of Abnormality. On the right side, a central donut chart provides a comprehensive overview of various abnormalities. The inner layer of the chart offers a detailed breakdown by gender, while the outer layer provides a nuanced breakdown by age group. b) The CPSC dataset is visualized similarly to the TNMG dataset, focusing on specific age groups and genders. c) The SNH dataset uses the same visual style as the TNMG dataset, incorporating organized data for age groups and genders.

The TNMG dataset, a comprehensive collection of ECG recordings, offers a valuable resource for investigating the prevalence of ECG abnormalities in the general population. Fig. 2a) presents a striking proportion of normal ECG records in the TNMG dataset, with over 80% of the total records displaying no labelled abnormality. This finding is consistent with the theoretical expectation of a healthy population, as demonstrated in Table 1. Moreover, the distribution of ECG abnormalities across different age groups in the TNMG data aligns with the known increased prevalence of abnormalities in the elderly population. Notably, the number of patients with abnormalities declines beyond 75, possibly due to lower survival rates among the elderly population with abnormalities. Overall, the present findings highlight the importance of utilizing large and diverse datasets such as TNMG to gain insights into the expected distribution of ECG abnormalities in the general population while also identifying age as a crucial factor in the prevalence of certain cardiac pathology.

The analysis of abnormality classifications within the dataset demonstrates a relatively uniform distribution, with each type of abnormality occurring at comparable frequencies. Furthermore, this consistent distribution pattern is maintained across both genders, with no apparent gender-based variations observed. The age-based breakdown of abnormality classifications is also consistent with the general trend seen in the overall breakdown of age groups within the dataset. Notably, most cases are concentrated within the middle age groups, underscoring the utility of this dataset for developing predictive models for abnormalities. These findings offer valuable insights into the prevalence and distribution of abnormalities in the general population and emphasize the importance of utilizing large and diverse datasets for developing robust predictive models for abnormality detection and classification.

#### China Physiological Signal Challenge 2018 (CPSC)

The CPSC training dataset is publicly accessible and includes an open-source collection of 6,877 12-lead ECG records. The recordings ranged from 6s to 60s and were sampled at a rate of 500 Hz [13]. The ECGs are divided into eight different types of abnormality, with AF, 1dAVb, LBBB, and RBBB overlapping with the TNMG dataset. Additionally, ST-segment depression (STD), premature atrial contraction (PAC), ST-segment elevation (STE) and premature ventricular contraction (PVC) are also represented.

In order to align the CPSC dataset with the TNMG dataset introduced earlier, patients diagnosed with STD, PAC, STE, and PVC in their first labelled condition are excluded from our analysis. The ECG tracings are also resampled to 400 Hz to ensure comparability with the TNMG dataset. The breakdown of the processed CPSC dataset is shown in Fig. 2b).

The dataset contains a high proportion of patients with abnormality, with approximately 75% of the total 4,622 unique ECG tracings exhibiting one or more types of abnormality. This high proportion is not a result of the reduction in dataset size but rather an inherent characteristic of the original dataset. The CPSC dataset follows general trends in the population with a higher frequency of abnormality among elderly patients. However, the distribution of patients across the four classifications of abnormality is uneven. The high proportion of patients with Right Bundle Branch Block (RBBB) does not reflect the population and makes the dataset uneven for training algorithms.

#### Shaoxing and Ningbo Hospitals (SNH)

The SNH dataset, open to the public, contains 45,152 12-lead ECG recordings gathered between 2013 and 2018 [25] [26]. These recordings are 10 seconds long, and a licensed physician has assigned labels to each one indicating its association with one or more of the 63 abnormality types. In order to conform with the TNMG dataset, only ECG tracings that are categorized as normal or fall under the six classes existing in TNMG have been retained. Consequently, the total number of recordings has been decreased to 34,033. Furthermore, all tracings have been resampled at a rate of 400 Hz. The processed SNH dataset is presented in Fig. 2c).

The SNH dataset highlights a significant prevalence of abnormalities among patients, with approximately 70% of the population presenting one or more types of abnormality. Upon closer examination of the patient classifications, the dataset reveals a significant skew towards SB, with more than half of the abnormality cases categorized as such. Such an imbalance towards SB in the dataset may pose a challenge when training an abnormality classification model.

### 3.2 Subsets

To investigate how training data characteristics affect the model’s performance and generalization ability, this study selected subsets of ECG tracings from the TNMG dataset, using different selection criteria. The same model was trained on each of the subsets, and a crossdataset performance analysis was conducted to examine the model’s performance and generalization capacity. The model is also compared to the model trained in [19].

By using different selection criteria for the training data subsets, this study aims to provide insights into which characteristics of the training data have the most significant impact on the model’s generalization ability. Furthermore, by analyzing the model’s cross-dataset performance, this study can assess the degree to which the model can apply what it learned from the training data to new datasets. The findings from this study can inform future work on developing models with robust generalization capabilities in ECG analysis.

### 3.3 Attention Mechanism

Attention mechanisms have been shown to improve the performance of models [3], however, this study goes beyond and examines the effect of attention mechanisms on generalization. In this study, an additional attention layer is proposed to be added to the base model, which will also be trained on the four subsets of ECG tracings. The performance of this model with the attention mechanism will be compared to the performance of the base model without the attention mechanism, and their cross-dataset generalization abilities will be evaluated.

The proposed base model with the attention mechanism is illustrated in Fig. 1b). By including an attention mechanism, the model can selectively focus on the most informative regions of the ECG tracings, which could potentially improve the model’s ability to generalize to new datasets. The findings from this study could provide insights into the benefits of incorporating attention mechanisms in ECG analysis models and how they impact the models’ generalization capabilities.

### 3.4 Evaluation Metrics

The models in this study are evaluated using the following performance metrics. In binary classification, TP (True Positive), TN (True Negative), FP (False Positive), and FN (False Negative) are defined, and *F*_1_ represents the harmonic mean of precision and recall, offering a balanced evaluation that accounts for both metrics.

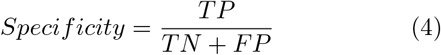

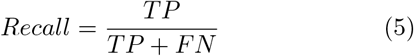

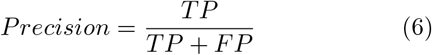

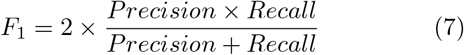

Due to the varying degrees of class imbalance observed in all three datasets, where one class significantly outweighs the other, the *F*_1_ score emerges as a more appropriate metric. By taking into account both true positives and false negatives, the *F*_1_ score offers better evaluation and sensitivity for imbalanced datasets.

A higher score typically indicates better performance of the model. While *F*_1_ score indicates the overall balanced performance of the model.

## 4 Experiment

### 4.1 Subsets

This study aims to investigate the influence of distinct selection criteria for training data on a proposed model architecture’s performance and generalization capacity. The model will be trained using four subsets of 12-lead ECG data from the TNMG dataset, each consisting of 21,000 traces, with varying selection criteria as presented in Table 2. The study’s main objective is to analyze how specific attributes or characteristics of the training data impact the performance and generalization ability of the model when applied to various datasets.

**Table 2:** Subset Selection Criteria.

| Subset | Selection Criteria |
| --- | --- |
| I | 3,000 recordings from each of the six ECG classification types and 3,000 normal readings |
| II | Same selection criteria as the approach I, but favour patients with multiple types of abnormality |
| III | Select the 1,500 oldest and youngest records for each ECG classification |
| IV | Randomly selected |

As a result of some patients having more than one type of abnormality, there may be repeated selection of patients from the TNMG dataset. The remaining data required to reach 21,000 records is randomly drawn from the dataset. These repeated selections may introduce discrepancies in the overall statistics. The statistics of the subsets are summarized in Table 3.

**Table 3:** Subsets statistics with charts displaying different age groups with distinct bars for females and males. The charts organize the female bars on the left and the male bars on the right for visual clarity.

| Subset | I | II | III | IV |
| --- | --- | --- | --- | --- |
| 1dAVb | 3,234 | 6,677 | 3,341 | 333 |
| RBBB | 4,320 | 7,020 | 3,819 | 589 |
| LBBB | 3,443 | 4,935 | 3,411 | 340 |
| SB | 3,367 | 3,624 | 3,324 | 357 |
| AF | 3,561 | 4,043 | 3,846 | 377 |
| ST | 3,299 | 3,219 | 3,395 | 455 |
| Normal | 3,000 | 3,000 | 3,000 | 18,793 |

### 4.2 Training

The architecture of the base model shown in Fig. 1 is trained on each of the subsets, and its performance is evaluated on a test dataset presented in [19]. The performance of these models is compared to that of a model trained in [19], and their cross-dataset generalization ability is assessed by testing the models on the CPSC and SNH datasets.

Similarly, we train the proposed model with the attention mechanism illustrated in Fig. 1b) on the subsets and select the best-performing model for evaluation of its cross-dataset generalization ability. The performance is compared to that of the base model trained in [19], and its generalization capacity is evaluated.

The hyperparameters previously used in [19] were deployed to train our model. These include 16 for kernel size, 64 for batch size, an initial learning rate of 0.001, the Adam optimizer, 0.8 for dropout rate, and epochs of 200 with a patience value of 9 epochs for early stopping. By utilizing these hyperparameters, we aim to ensure consistency in our experiments and facilitate direct comparison with prior research. Such an approach enables a more rigorous evaluation of our proposed model architecture’s performance in ECG signal classification tasks.

### 4.3 Model Testing

The models were evaluated on the test dataset described in [19], which consists of 827 unique 12-lead tracings of ECG records. Further details regarding the breakdown of the test dataset can be found in Table 4.

**Table 4:** TNMG test-set statistics by age groups with separate bars for females and males. The female bars are positioned on the left, and the male bars on the right for visual clarity.

| Type | Count |
| --- | --- |
| 1dAVb | 28 |
| RBBB | 34 |
| LBBB | 30 |
| SB | 16 |
| AF | 13 |
| ST | 36 |
| Age |  |
| 90+ |  |
| 75-89 |  |
| 60-74 |  |
| 45-59 |  |
| 30-44 |  |
| 15-29 |  |

To assess the generalization capacity of the models across different datasets, tests of the models were conducted on the CPSC and SNH datasets. The performance of the models was evaluated using the proposed metrics, and the results were analysed to determine their generalization performance.

## 5 Result

The experiment results are divided into two subsections for presentation. In the first subsection, we focus on the findings related to the base model architecture, exploring how its performance varies based on different characteristics of the subsets used in training. In the second subsection, we shift our attention to the proposed model with attention mechanisms evaluating its performance across the same subsets. By segregating the analysis, we can understand both the base model’s behavior and the potential enhancements brought about by incorporating attention mechanisms.

### 5.1 Effect of Dataset Characteristics

The models trained on the subsets are tested on the testset of [19] and are compared to the result of the trained model in [19]. The performance metrics of the models are presented in Table 5.

**Table 5:** Analyzing DNN Model Performance: Evaluating Inference Results on TNMG test set, CPSC, and SNH Datasets for Various Subsets (I, II, III & IV) versus the Full Dataset Training.

| Subset | I | II | III | IV | Full[19] |
| --- | --- | --- | --- | --- | --- |
| **Inference on TMNG Test-set** |  |  |  |  |  |
| Precision | 0.81 | 0.77 | 0.80 | 0.79 | 0.92 |
| Recall | 0.87 | 0.93 | 0.89 | 0.82 | 0.94 |
| Specificity | 0.99 | 0.99 | 0.99 | 0.99 | 1.00 |
| F1 Score | 0.84 | 0.83 | 0.83 | 0.80 | 0.93 |
| **Inference on CPSC Dataset** |  |  |  |  |  |
| Precision | 0.93 | 0.88 | 0.90 | 0.80 | 0.91 |
| Recall | 0.84 | 0.84 | 0.86 | 0.76 | 0.84 |
| Specificity | 0.98 | 0.97 | 0.98 | 0.95 | 0.98 |
| F1 Score | 0.88 | 0.86 | 0.88 | 0.77 | 0.87 |
| **Inference on SNH Dataset** |  |  |  |  |  |
| Precision | 0.48 | 0.51 | 0.48 | 0.15 | 0.48 |
| Recall | 0.50 | 0.55 | 0.53 | 1.00 | 0.53 |
| Specificity | 0.85 | 0.82 | 0.82 | 0.00 | 0.82 |
| F1 Score | 0.47 | 0.50 | 0.49 | 0.22 | 0.48 |

The trained models underwent evaluation on the CPSC dataset through model inference, and their performance was compared to that of the model presented in [19]. The results are summarized in Table 5.

The models were ultimately inferred on the SNH dataset, and their effectiveness was compared to that of the model introduced in [19]. The comparison results are illustrated in Table 5.

### 5.2 Boosted Model with Attention Mechanism

To comprehend the impact of the attention mechanism on the generalizability of the model, we trained the proposed model architecture depicted in Fig. 1b) on the four subsets. Its performance on the test-set of [19] is presented in Table 6.

**Table 6:**
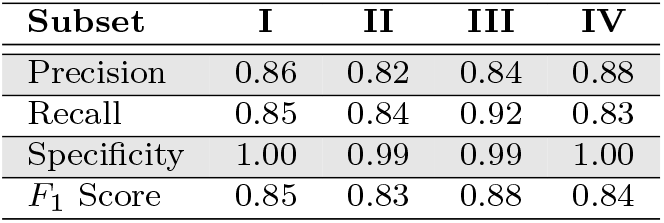
The inference results of the Attention-DNN model trained on different subsets (I, II, III & IV) evaluated on the TNMG test set.

| Subset | I | II | III | IV |
| --- | --- | --- | --- | --- |
| Precision | 0.86 | 0.82 | 0.84 | 0.88 |
| Recall | 0.85 | 0.84 | 0.92 | 0.83 |
| Specificity | 1.00 | 0.99 | 0.99 | 1.00 |
| $F_1$ Score | 0.85 | 0.83 | 0.88 | 0.84 |

The top-performing model, which incorporates the attention mechanism and achieves the highest *F*_1_ score (Subset III), was evaluated on both the CPSC and SNH datasets through model inference. Its performance was compared to that of the TNMG model introduced in [19], and the results are presented in Table 7.

**Table 7:** Comparative inference evaluation of model performance trained on TNMG subset III to the original DNN model trained on the entire TNMG dataset.

|  | **Attention DNN III** |  | **DNN[19]** |  |
| --- | --- | --- | --- | --- |
| Dataset | CPSC | SNH | CPSC | SNH |
| Precision | 0.90 | 0.50 | 0.91 | 0.48 |
| Recall | 0.86 | 0.57 | 0.84 | 0.53 |
| Specificity | 0.97 | 0.81 | 0.98 | 0.82 |
| $F_1$ Score | 0.88 | 0.52 | 0.87 | 0.48 |

## 6 Discussion

Based on the experimental results, it is evident that the training data’s characteristics significantly impact the model’s performance, even when using the same model structure and amount of training data. The models trained on subsets I, II, and III exhibited better performance compared to the model trained on subset IV, achieving *F*_1_ scores of 0.84, 0.83, 0.83, and 0.80, respectively. The improved performance of subsets I, II, and III can be attributed to the balanced nature of their training datasets, which had similar abnormality classifications. While a larger dataset can lead to better in-sample performance, it is crucial to acknowledge that this performance boost may be attributed to the presence of data from each class of abnormality. However, considering that the subset comprises only around 1% of the original dataset, the models trained on these subsets still demonstrate commendable performance.

The story changes when it comes to model generalization. As discussed earlier, for inference performance on the CPSC dataset, models trained on subsets I, II, and III still outperformed the model trained on subset IV, demonstrating the significance of a balanced dataset, achieving *F*_1_ scores of 0.88, 0.86, 0.88, and 0.77, respectively. However, what is even more surprising is that Ribeiro *et al*.’s model, which was trained on the entire TNMG dataset and achieved an *F*_1_ score of 0.87, did not perform better than the models trained on smaller subsets I and III. It is only marginally better performing than the model trained on subset II. This discovery is particularly interesting, considering that the subsets use only approximately 1% of the entire TNMG dataset.

This finding suggests that a well-balanced and properly curated smaller dataset can still lead to competitive model performance compared to larger and more diverse datasets.

For inference performance on the SNH dataset, the model’s performance experienced a significant decrease, which aligns with the long-standing issue in biomedical machine learning models. These models often struggle to generalize well when faced with changes in data, equipment, environment, and patient characteristics. To address this problem, some researchers have proposed patient-specific models, and there is a growing body of research in this direction.

The Ribeiro *et al*. model achieved a considerably lower F1 score of 0.48 on the SNH dataset, indicating that even with a large dataset, it still faces challenges with model generalization in the biomedical sector. Conversely, the models trained on subsets I, II, and III outperformed those trained on subset IV, with F1 scores of 0.47, 0.50, 0.49, and 0.22, respectively. This reaffirms the importance of having a balanced dataset for training. Remarkably, the same interesting discovery was made when compared to Ribeiro *et al*.’s model on the SNH dataset. The model once again failed to outperform models trained on only 1% of its data. It performed lower than models trained on subsets II and III and slightly better than those trained on subset I. This discovery further confirms the previous findings observed in the CPSC dataset inference results. It underscores the importance of dataset curation and balance even when dealing with large datasets in the biomedical domain.

The final part of the experiment focuses on the impact of the attention mechanism on performance and generalization. The results clearly indicate that the attention mechanism enhances the performance of almost all subsets in terms of the *F*_1_ score, with a particularly significant improvement observed in subset IV, which initially had the lowest *F*_1_ score when tested the models on the TNMG test-set. The performance of the models has improved to *F*_1_ scores of 0.85, 0.83, 0.88, and 0.84, respectively, as shown in Table 6. This trend also translates into generalization, as evidenced by the comparison of Ribeiro *et al*.’s model and the model with the attention mechanism trained on subset III. The new model has outperformed Ribeiro *et al*.’s model in inference performance on both CPSC and SNH datasets illustrated in Table 7. Considering the fact that the new model was trained with only 1% of the TNMG dataset, this again confirms the previous finding of the importance of balanced data and confirms the positive impact of the attention mechanism on improving the model’s accuracy and inference performances.

Again, looking at the performance of the models trained on subsets, the performance between models trained on I, II and III are not clearly higher or lower, indicating that the selection criteria used for subset I, II, and III with age preference and multi-abnormality patients do not have a clear impact on the model performance. Therefore, balanced data is the most important factor. This study also clears many researchers’ doubts, encouraging using datasets with characteristics matching the general population. The TNMG dataset, claimed to be the largest ECG dataset, matches the proportion of various abnormalities in the general population. Nonetheless, models trained using this dataset struggle to exhibit the same level of generalization as models trained on a smaller dataset that is balanced. Therefore, researchers should use balanced ECG datasets for training to achieve the best-performing models.

## 7 Conclusion

The study emphasizes the crucial role of dataset characteristics in influencing model performance and generalization. Specifically, the researchers found that models trained on balanced subsets consistently outperformed those trained on unbalanced data, regardless of the latter’s larger size. This highlights the critical importance of dataset balance in achieving better model performance.

Moreover, introducing the attention mechanism proved to be a valuable enhancement, boosting model performance across various subsets. Notably, the most significant enhancement was observed in subset IV, which initially had the lowest performance. The positive impact of the attention mechanism was evident not only on the TNMG test-set but also on external datasets (CPSC and SNH), demonstrating its efficacy in improving model accuracy and generalization.

These findings hold significant implications for researchers and practitioners in the biomedical field. It strongly encourages using carefully curated, balanced datasets and adopting attention mechanisms to achieve the best-performing models. By addressing these factors, the research aims to contribute to advancing biomedical machine learning, ultimately leading to better healthcare outcomes in real-world settings.

## Data Availability

All data produced in the present study are available upon reasonable request to the authors

## 8 Acknowledgement

Zhaojing Huang would like to acknowledge the support of the Research Training Program (RTP) provided by the Australian Government.

## 9 Code Availability

To obtain the code used in this paper, request it from the corresponding author, but be aware of any conditions or restrictions on its use.

## 10 Data Availability

This paper employs publicly accessible datasets such as CPSC, SNH, and the TNMG dataset. The TNMG dataset is not publicly available, but access can be granted upon request via the owner’s approval.

## 11 Declaration of No Conflicts of Interest

The authors affirm that they have no conflicts of interest, including both financial and non-financial, to report.

